# A lightweight, automated neural network-based stage-specific malaria detection software using dimension reduction: The malaria stage classifier

**DOI:** 10.1101/2022.11.28.22282777

**Authors:** Preißinger Katharina, Kézsmárki István, Török János

## Abstract

Due to climate change and the COVID-19 pandemic, the number of malaria cases and deaths increased between 2019 and 2020 [1]. Reversing this trend and eliminating malaria worldwide requires improvements in malaria diagnosis, in which artificial intelligence (AI) has recently been demonstrated to have a great potential. Here, we describe an AI-based approach that boosts the performance of light (LM), atomic force (AFM) and fluorescence microscopy (FM)-based malaria diagnosis. As the main challenge, the stage-specific recognition of infected red blood cells (RBCs) usually requires large sets of microscopy images for training a neural network, which is difficult to obtain. Our tool, the Malaria Stage Classifier, provides a fast, high-accuracy recognition that works even with limited training sets due to a smart reduction of data dimension. Individual RBCs are extracted from an image, reduced to characteristic one-dimensional cross-sections, and classified. We show that our method is applicable to images recorded by various microscopy techniques. It is available as a software package at https://github.com/KatharinaPreissinger/Malaria_stage_classifier and can be used within a python environment. Technical support is provided by the corresponding author.

**Author summary:** The Malaria Stage Classifier is a software helping the user to detect and stage RBCs infected with malaria. Accurate recognition of malaria infected RBCs still imposes a challenge in endemic regions, as it is time-consuming and subjective. These deficiencies can be overcome by autonomous computer assisted recognition using neural networks (NNs). The Malaria Stage Classifier offers a user-friendly interface for the stage-specific classification of malaria infected RBCs into four categories—healthy ones and three classes of infected ones according to the parasite age. The use of data reduction, which forms the central element of the Malaria Stage Classifier, allows for a fast and accurate classification of RBCs. It is applicable for light, atomic force, and fluorescence microscopy images and allows for retraining the implemented NN with new images. Our simple concept further has the potential to be generalised for the classification of other cells or objects.

## Introduction

More ancient than human, malaria has caused millions of deaths until today [1]. Nowadays, the most common technique used worldwide is light microscopy of Giemsa-stained blood smears, which is time-consuming and heavily relies on human performance. To overcome these issues, computer-aided recognition of malaria-infected red blood cells (RBCs) has recently gained high attention.

As the number of cases is increasing due to the ongoing COVID-19 pandemic and climate change, the need for rapid and easily accessible recognition of the disease is rising. The infection is caused by five types of the *Plasmodium* genus, upon which *Plasmodium falciparum* causes the most deaths. Injected into the human body by a mosquito bite, the malaria parasites migrate to the liver cells, where they begin a phase of asexual reproduction, which is followed by the release of many thousands of merozoites. During their 48 h cycle in the blood stream, the parasites invade healthy RBCs and mature through three main stages—the ring, the trophozoite, and the schizont stage—, while altering the morphological and optical properties of the host RBCs [5, 6].

These well-explored transformations of the infected RBCs provide the basis for neural network (NN) based stage-specific recognition of malaria-infected RBCs in blood smears [7–11]. While clearly demonstrating the potential of NNs in malaria diagnosis, these pioneering approaches still face problems, e.g., in terms of sensitivity to different malaria species and stages. The following studies illustrate the state-of-the-art in this field.

Based on recent advances in high-resolution imaging techniques applied for the analysis of malaria-infected RBCs, especially topographic imaging [5, 12, 13] and infrared nano-imaging [14], recent studies on unstained RBCs [6] have demonstrated the applicability of NNs to analyse not only light microscopy images but also images recorded with other microscopy techniques. These works imply that, when combined with NN-based analysis, malaria diagnosis may be extended to other imaging methods. One of the main reasons for the use of NNs is the time saving through automatising the process and the elimination of human error, when working with big amounts of data [15]. The majority of the algorithms developed for the analysis of malaria-infected RBCs up to now are limited to Giemsa-stained images and rely on a two-stage recognition, sorting healthy RBCs from infected ones, followed by a stage-specific categorisation of the infected cells [16–19]. These algorithms have reached performances of *∼*80-98% with little or no pre-processing of microscopy images [20–23]. By adding characteristic attributes of the RBCs, including the colour scheme [17], the morphology of the RBCs [18], and their other statistical features [19] to the analysis, similar results have been achieved.

When working with two-dimensional data images, the number of parameters, that has to be fitted by the NN for the classification of RBCs, is extremely high, which strongly influences the performance of the network. This issue can be solved by reducing the dimension of the input data and may even improve the classification [24]. To handle the loss of data that comes with the reduction of the dimension, features can be selected, which capture the characteristic properties of the RBCs [18, 19, 25].

The complicated handling of malaria culturing and sample preparation introduces some limitations to the application of NNs, which typically results in a small training set of a few thousands of images and an uneven distribution of RBC categories. The latter leads to overfitting [26], significantly influencing the performance of the network. One way out of this dilemma is the use of data augmentation, which increases the number of images and equalises the distribution of healthy and infected RBCs [20, 21], thus improving the performance of the classification [22, 23].

### Design and Implementation

The Malaria Stage Classifier is designed to facilitate and accelerate the staging of malaria infected RBCs in microscopy images. Due to its robustness against imaging platform-specific features, it is applicable to a wide range of light microscopy images. The interface of the application is arranged in tabs, which makes it easy to follow the image processing steps. The Malaria Stage Classifier further offers the possibility to manually optimise the cell detection and classification.

### Data loading

From the microscopy measurement, the algorithm receives two kinds of input, text files from atomic force microscopy and images from fluorescence and light microscopy. Both inputs are treated as matrices and converted to greyscale for further processing. In case of atomic force and some light and fluorescence microscopy images, the contrast between background and RBCs is not strong enough to locate single cells, which impacts the accuracy of the detection. Hence, the images are binarised based on pixel intensities by Otsu’s method [27]. While the processing of atomic force microscopy images requires an additional step, it is sufficient to enhance brightness, sharpness, and contrast in the light and fluorescence microscopy images. Python offers a module for automatic enhancement of images by a manually chosen factor [28], which can be applied to highlight the RBCs in contrast to the image background. The processed images are then used for cell detection, employing the Hough gradient method. While the detection parameters are preset, they can be manually adjusted by the user.

### Reduction of dimensionality as a tool for feature selection in malaria-infected RBCs

The accurate detection of the malaria blood stage plays a crucial role in diagnosis. Therefore, we tried to find a suitable measure for the characteristic features of the intra-erythrocytic stages, which should be insensitive to external noise, often present in AFM and fluorescence microscopy images due to tip contamination or background illumination. We determined characteristic cuts through RBC images by two measures: the geometric centre and the centre of gravity, which are defined by:

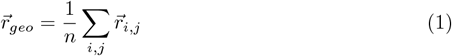

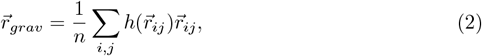

where 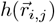 denotes the height (AFM) or intensity (fluorescence and light microscopy) of a pixel at position 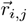. The coordinates inside the RBCs are calculated form the circle fit of the contour, which is returned by the Hough gradient method. The idea behind this approach is to localise the parasites inside the cell, i.e. the coordinates are weighted by their corresponding height or intensity values. Fig 1 shows the effect of a region with lower intensity on two images of cylinders. The presence of the hole inside the cylinders leads to a shift of the gravitational centre, either away or towards the hole, which is indicated by an arrow. In case of the dark cylinder, the shift is higher, as the image values of black pixels are 0 and the values of white pixels 255. Therefore, the presence of the white hole has a larger effect on the position of the gravitational centre. Similarly, this shift can be observed for RBC images. From this follows that the height or intensity values along the straight line through both centres represent a cross-sections of the parasite. In the further course of the paper, this straight is defined as the parasite cut.

**Figure 1.**
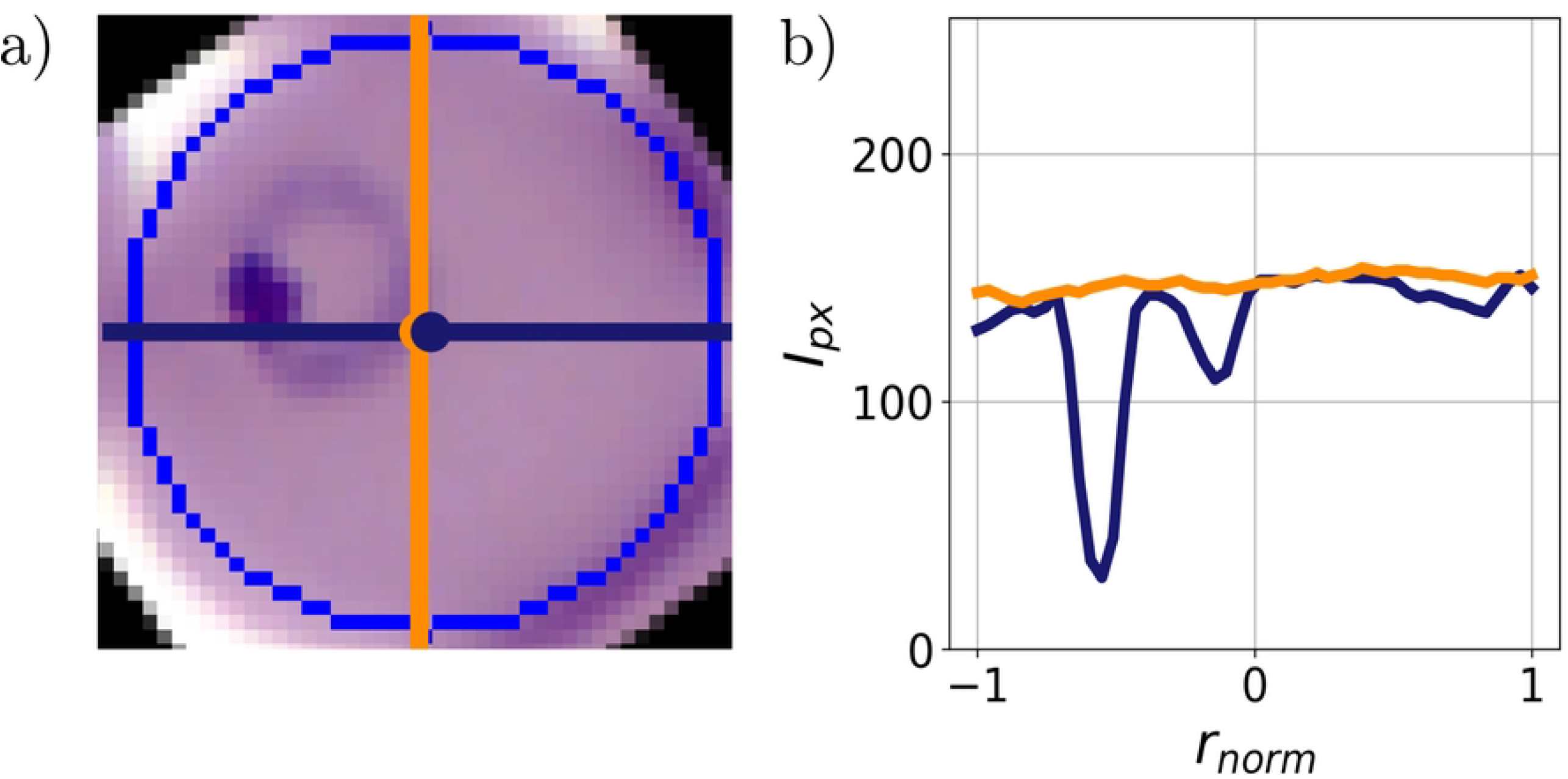
Location of the geometric and gravitational centre in simple cylinders with holes. Geometric and gravitational centre are shown as an orange and blue cross. The effect of the hole on the location of the gravitational centre is shown for a black (a) and white cylinder.

An evaluation of the efficiency of the presented measure is shown in our paper about the stage-specific detection of malaria-infected RBCs based on dimension reduction [3]. The efficiency was tested on at least 350 ring, 550 trophozoite, and 550 schizont-stage parasites. In more than 50% of the cases, the parasite cut captures the ring-stage parasite, while the detection probability of the trophozoite-stage parasite is > 85%. The schizont-stage parasites typically fill up most of the RBC, thus characteristic cuts going through the always intersect the parasite. To fully represent the characteristic features of single cells, the parasite cut is supplemented by an additional cut, spanning 90^*o*^ with it. A representative image is shown in Fig 2.

**Figure 2.**
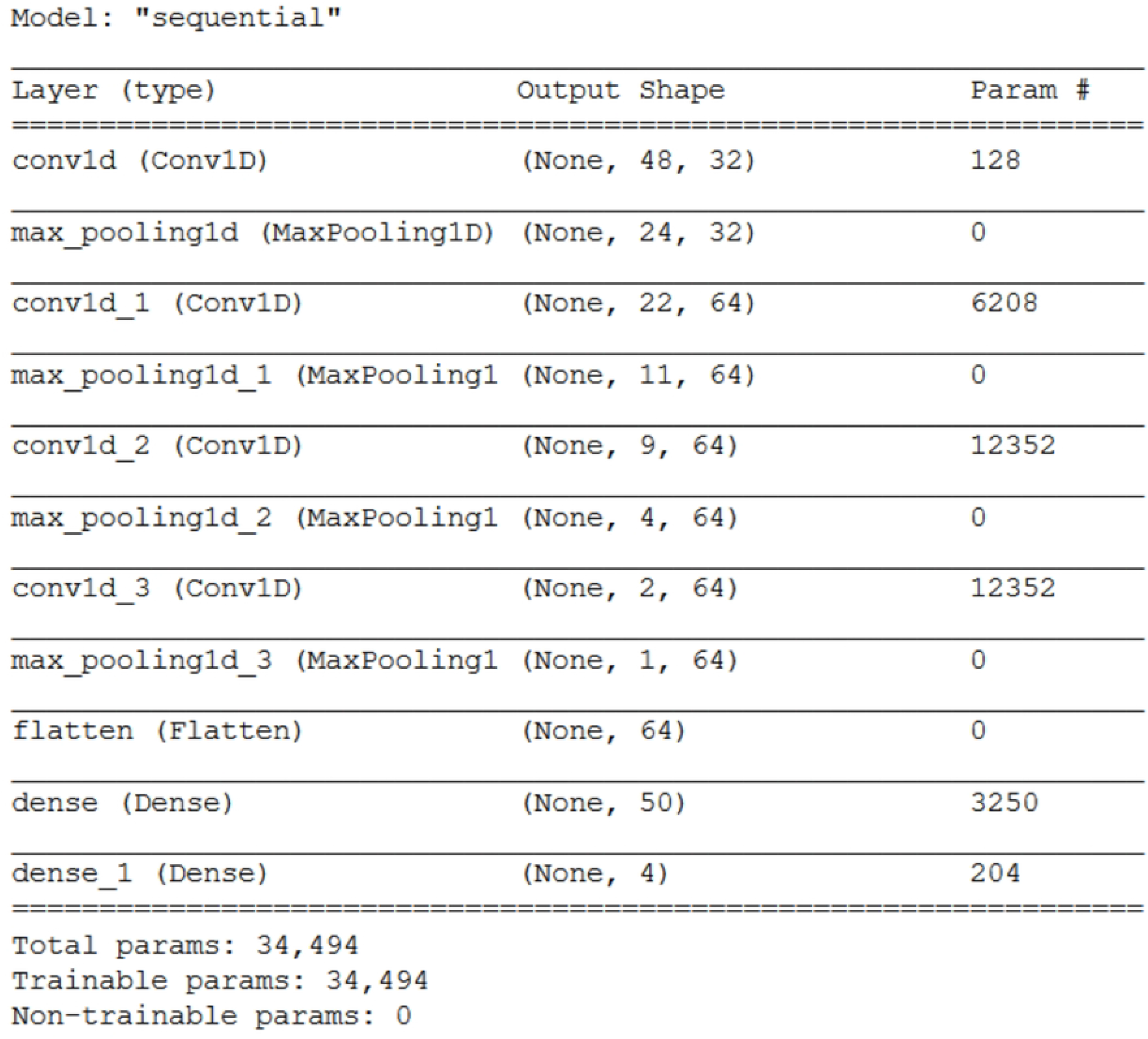
Characteristic cuts of a parasite. (a) Light microscopy image showing the parasite cut (blue) and the additional cut spanning 90^*o*^ (orange) with it. (b) Corresponding intensity profiles. The contour of the cell is shown in blue.

### Neural network architecture

When dealing with images, the most suitable models are convolutional neural networks (CNN). Generally, they contain convolutional, pooling, and dense layers [31], which we combined in such a way as to fulfil the following conditions: applicability to RBC images and stage-specific cuts, classification into four categories: healthy, ring, trophozoite, schizont, minimum performance of > 90%, smallest possible amount of fit parameters to guarantee short computation time. The models were tested on a set of malaria-infected RBC images, recorded by AFM and balanced with data augmentation by rotation. For a representative result, all models were run for 20 epochs. In each model, the convolutional layers are followed by a pooling layer and two fully-connected layers at the end of the network. As an option, the user can retrain the pre-trained networks provided by the application with new images.

### Post-computation

After the classification of the microscopy images, the results can be saved as a csv or text-file, returning the amount and ratio of healthy RBCs and intra-erythrocytic stages.

### Implementation

The Malaria Stage Classifier is accessible as a GUI, which is arranged in tabs. It has been developed using Python 3.7 with the following dependencies: *numpy* [34] and *pandas* [35] for the data analysis. For the cell detection, *OpenCV* [36] together with *skimage* [37] and *matplotlib 3*.*5*.*2* [38] for the data visualisation are needed. To integrate the pre-trained NN, *tensorflow* [39] with keras has to be included in the algorithm. The interface further requires *tkinter* [40] and the libraries *os, sys, csv, traceback* for handling errors and the output files, as well as the library *webbrowser* to open system folders and external links.

Common errors are prevented by messages, detailing the problem and providing a solution. Furthermore, the errors saved in a log file to allow for bug-fixing by any developer. While the user is able to modify a few parameters through the GUI, suitable values are suggested for each case and all parameters can be set back to the default values.

## Results and discussion

In the previous sections, we have described the methods, which form the two main parts of the “Malaria Stage Classifier”. To find suitable parameters for the presented methods, we tested the cell detection algorithm and the performance of the NNs for all microscopy techniques.

### Segmenting and image processing

The RBCs in the microscopy images are detected in three steps: localising edges, finding the centre of the object, and calculating its radius [29]. The method is controlled by five parameters, mDist, par1, par2, minR, and maxR. mDist defines the minimum pixel distance between the centres of two objects. par1 is the threshold value for edge detection by the Canny edge detector [30]. par2 sets the threshold for the number of edge points to declare the object a circle. minR and maxR set the minimum and maximum size of the radius in pixels. The optimum values for each parameter are shown in table 1, which allow for the localisation of more than 95% of all RBCs (approx. 7000 for each imaging technique). We note that limitations can occur for not circular or overlapping RBCs, which can be handled by manually adjusting the detection parameters. The centre and radius of the cells, detected by the Hough gradient method, define the contour and image coordinates associated with the RBCs, which are used for the calculation of the geometric and gravitational centre. While the optimum values work for the tested images, we added the option to manually adjust each parameter in order to improve the cell detection. Fig 3 shows a representative microscopy image after running the cell detection algorithm with the default parameters.

**Table 1.** Optimum values for the sensitivity of the Hough gradient method for various imaging methods with the probability of cell detection.

| Imaging method | *mDist*<br>[px] | *par1*<br>[px value] | *par2*<br>[px value] | *minR*<br>[px] | *maxR*<br>[px] | Detection probability |
| --- | --- | --- | --- | --- | --- | --- |
| LM | 28 | 20 | 18 | 14 | 30 | 95% |
| AFM | 28 | 10 | 8 | 14 | 30 | 96% |
| FM | 28 | 20 | 18 | 14 | 30 | 97% |

**Figure 3.**
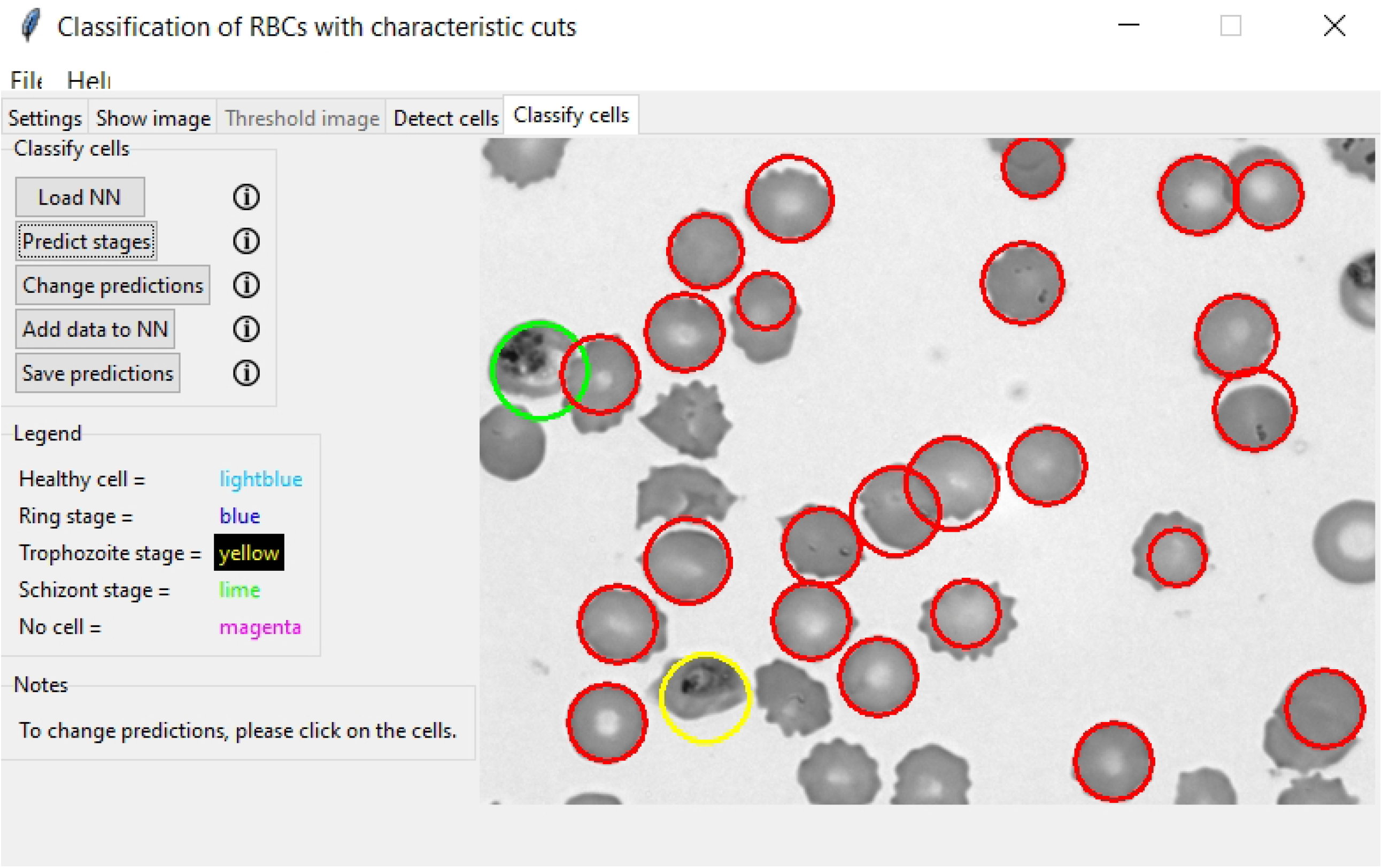
Cell detection in light microscopy image. The detected cells are marked with a cyan circle.

### Choosing the appropriate network

In order to find a suitable network for the classification of RBCs, we tested various network architectures on a set of RBC images. In table 2, the influence of the amount of feature maps is shown on a selection of the tested architectures. Model M4 provides the best compromise between the amount of fit parameters and classification accuracy. As the network was prone to overfitting, we added the kernel regulariser L2 with a hyperparameter of 0.001 to the first fully-connected layer. The complete architecture of the NN for a one-dimensional input, which is used for the classification of the single RBC in microscopy images, is given in Fig 4.

**Table 2.** Classification accuracy on a test set of RBCs images for various network architectures.

| Network | feature map | performance | parameters |
| --- | --- | --- | --- |
| M1 | $4 \times 32$ | 80% | 81566 |
| M2 | $2 \times 32, 2 \times 64$ | 87% | 68446 |
| M3 | $4 \times 64$ | 92% | 114878 |
| M4 | $1 \times 32, 3 \times 64$ | 96% | 96126 |

**Figure 4.**
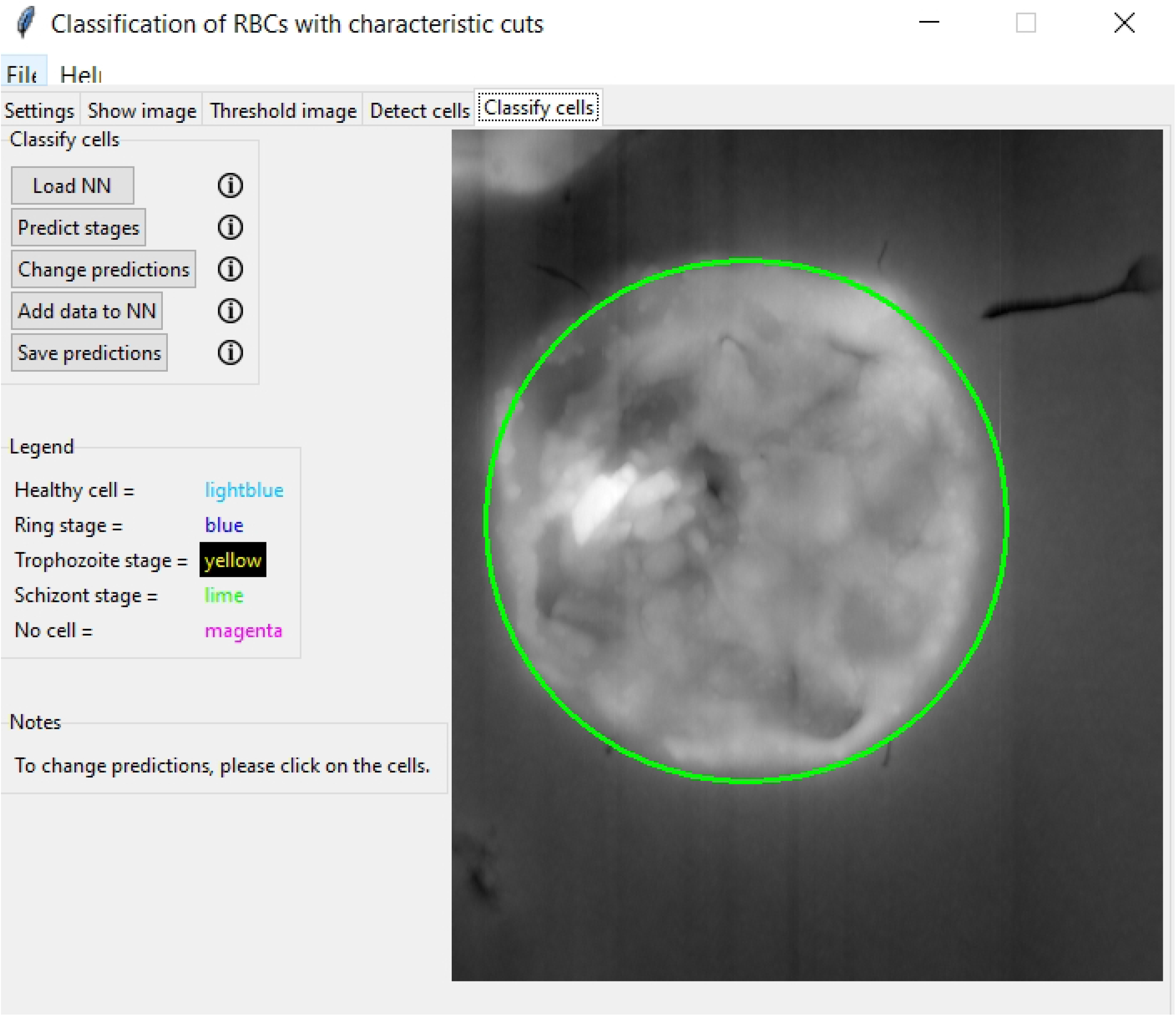
Neural network architecture. The network is used for the stage-specific classification of RBCs.

Our results show that network M4 performs best on the test data, reaching 96% accuracy. Therefore, we chose this architecture to test the classification accuracy in dependence of the imaging techniques.

### Performance of the neural network on the characteristic cuts

To predict the intra-erythrocytic stages of malaria based on the characteristic cuts, our convolutional neural network was trained on approx. 70000 and tested on approx. 8000 sets of characteristic cuts, for each imaging method respectively. The training and test set for light microscopy contains RBC images of 17 malaria-infected patients from data published by Abbas et *al*. [32]. The results are shown in Fig 6 in form of a confusion matrix. For multi-class problems, precision and recall are defined as

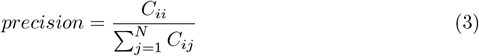

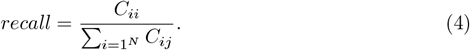

The overall accuracy of the classification is calculated as the ratio of correct predictions, independent of the categories, to the total of values in the confusion matrix [33].

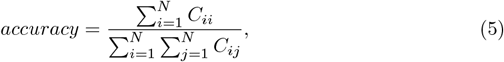

where *N* the number of classes, *i* the row index and *j* the column index. Fig 6 shows the overall performance of our network on the test data. With the characteristic cuts as input, it reaches more than 98% on the test sets, irrespective of the imaging method. Together with the cell detection algorithm, the pre-trained networks presented in the figure form the main elements of the Malaria Stage Classifier. The successful classification of infected RBCs is shown in Fig 5, where the healthy cells are surrounded by a red circle and the infected cells by a yellow and green circle, denoting the intra-erythrocytic stages trophozoite and schizont.

**Figure 5.**
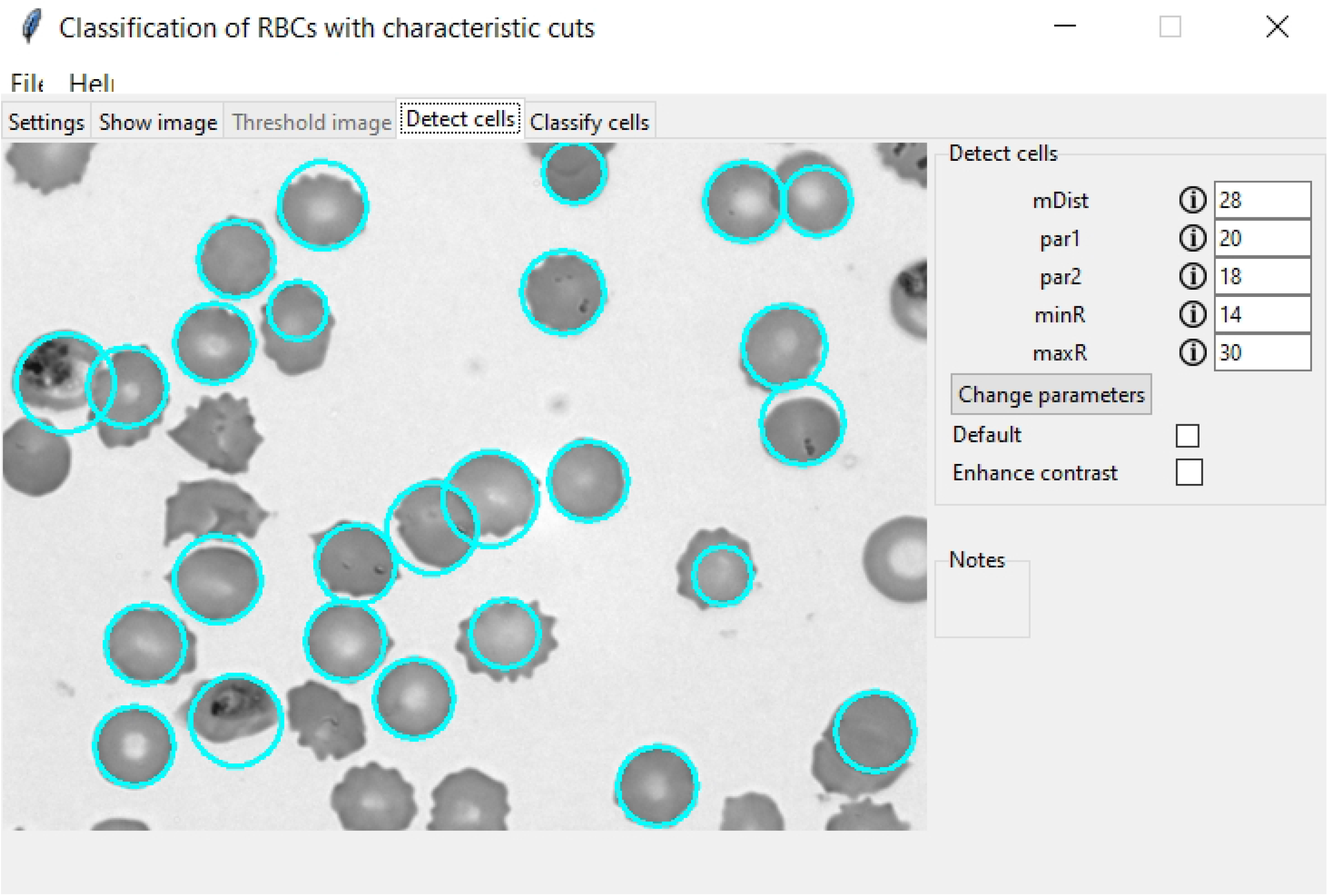
Classification of RBCs in a thin blood film. The image shows a thin blood film of a malaria-infected patient [32].

**Figure 6.**
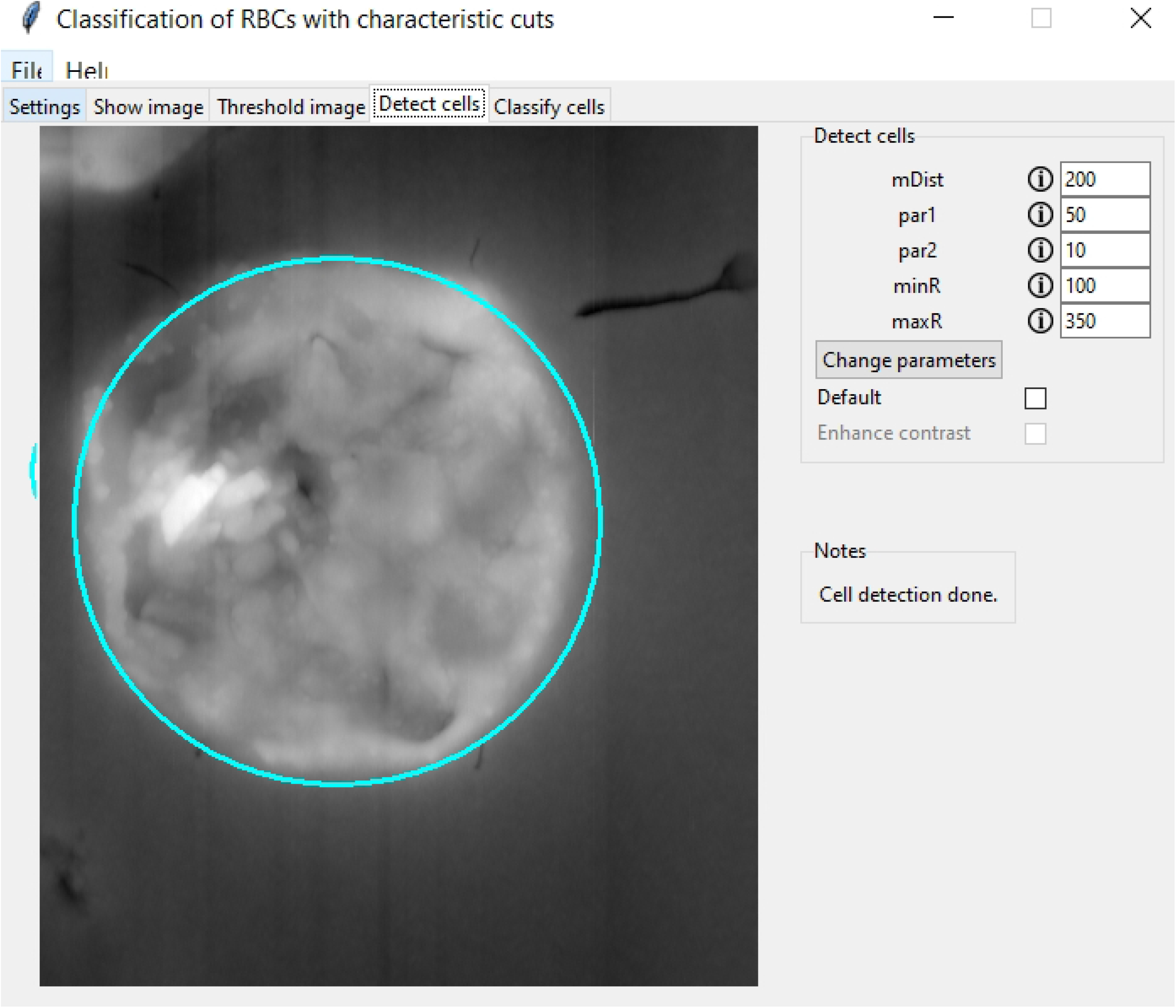
Performance of the NN on characteristic cross-sections. The classification results on the test set, as obtained on the characteristic cuts through light microscopy (black), AFM (blue) and fluorescence microscopy (red) images, are respectively summarised in the confusion matrix. The labels of the rows are the categories predicted by the NN-based classifier, while the labels of the columns indicate the classification by human experts. The diagonal elements show the correctly predicted cells, while the off-diagonal elements correspond to false classifications. The last column shows the precision for each category and the recall is shown in the bottom row. Each field displays the number of counts and the corresponding percentage with respect to the total number of cells in the test set. The overall classification performance is displayed in the grey field in the bottom right corner.

Here, we introduce a simple cell detection algorithm to classify malaria-infected RBCs in microscopy images into four categories: healthy, ring-, trophozoite-, and schizont-stage parasites. Our approach is not limited to a certain microscopy technique but demonstrated to be universally applicable for fundamentally different imaging techniques, which was demonstrated via the high classification accuracy on images recorded with Giemsa-stained light microscopy, AFM, and fluorescence microscopy. The reduction of dimension by selecting characteristic features of the RBCs significantly boosts the speed and classification accuracy of the network. This simple concept can easily be applied for the classification of general objects. Given the rising number of techniques, successfully applied for the imaging of RBCs, the algorithm can be augmented for any method with high contrast, formatted as text or image file.

## Availability and Future Directions

The “Malaria Stage Classifier” is deposited at the git hub repository https://github.com/KatharinaPreissinger/Malaria_stage_classifier or the archive https://zenodo.org/record/7261800. All details about the execution and usage of the package are documented in the documentation of our repository on https://github.com/KatharinaPreissinger/Malaria_stage_classifier or on https://malaria-stage-classifier.readthedocs.io/en/latest/index.html. There, we provide a tutorial for downloading the package and files with test images. The data set required for re-training the NN can be downloaded from https://zenodo.org/record/6866337.

Researchers are encouraged to implement new methods themselves, as the documentation is easily understandable and can be found at https://malaria-stage-classifier.readthedocs.io/en/latest/index.html. In case of low contrast microscopy images, the option of contrast improvement can be implemented, including thresholding for text files and the enhancement of brightness and contrast for image files. Moreover, we provide the opportunity to increase the data set, which is used to train the NN, to improve the classification accuracy of the algorithm. After loading the NN and visually verifying the accuracy of the classifications, the researchers have the possibility to add the characteristic single-RBC cross-sections to the original images and to retrain the NN with the new data set. The dataset for the new training is deposited at https://zenodo.org/record/6866337.

## Supporting information

### User manual

The package is available open source on git hub repository https://github.com/KatharinaPreissinger/Malaria_stage_classifier or the archive https://zenodo.org/record/7261800. In the following, the program structure is explained on a sample atomic force microscopy image. The interface of the package is built with five tabs, where each performs one step of the stage-specific classification of RBCs. Starting with the general settings, the algorithm only allows text or image files as input. Depending on the file type, the user can set the number of header lines the input text file contains. In case of the sample image, the value is set to three and confirmed by the button “Set file format”. As the program is designed for multiple imaging methods, the respective technique determines the possible actions on the input file. The settings are completed by selecting an output path and the output file type, see Fig 7.

**Figure 7.**
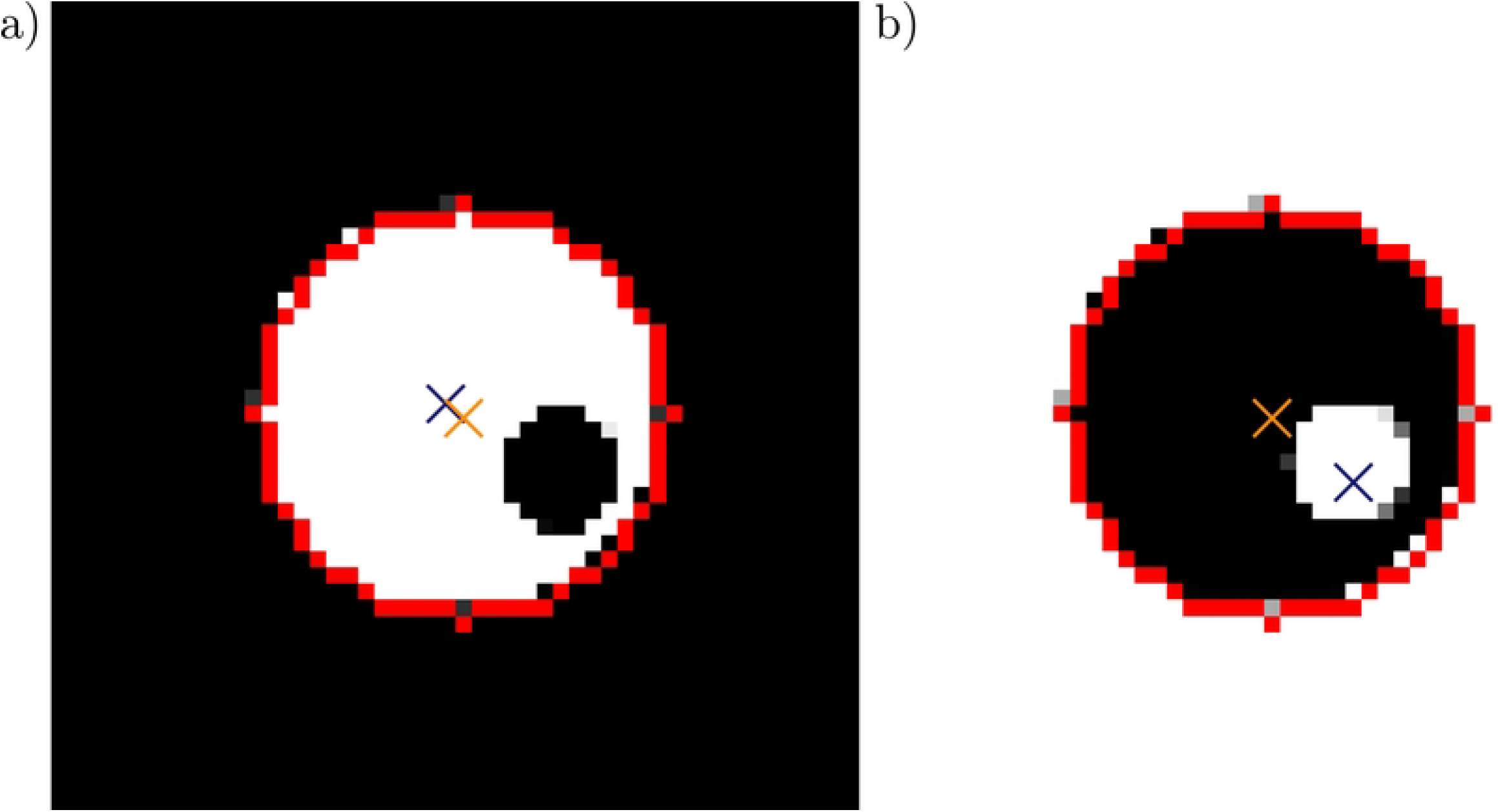
Setting of initial parameters: input image, its format, imaging method, memory location of the results

In the second tab, the input image is displayed to show the location and stage of the cells. A screenshot of the interface is presented in Fig 8.

**Figure 8.**
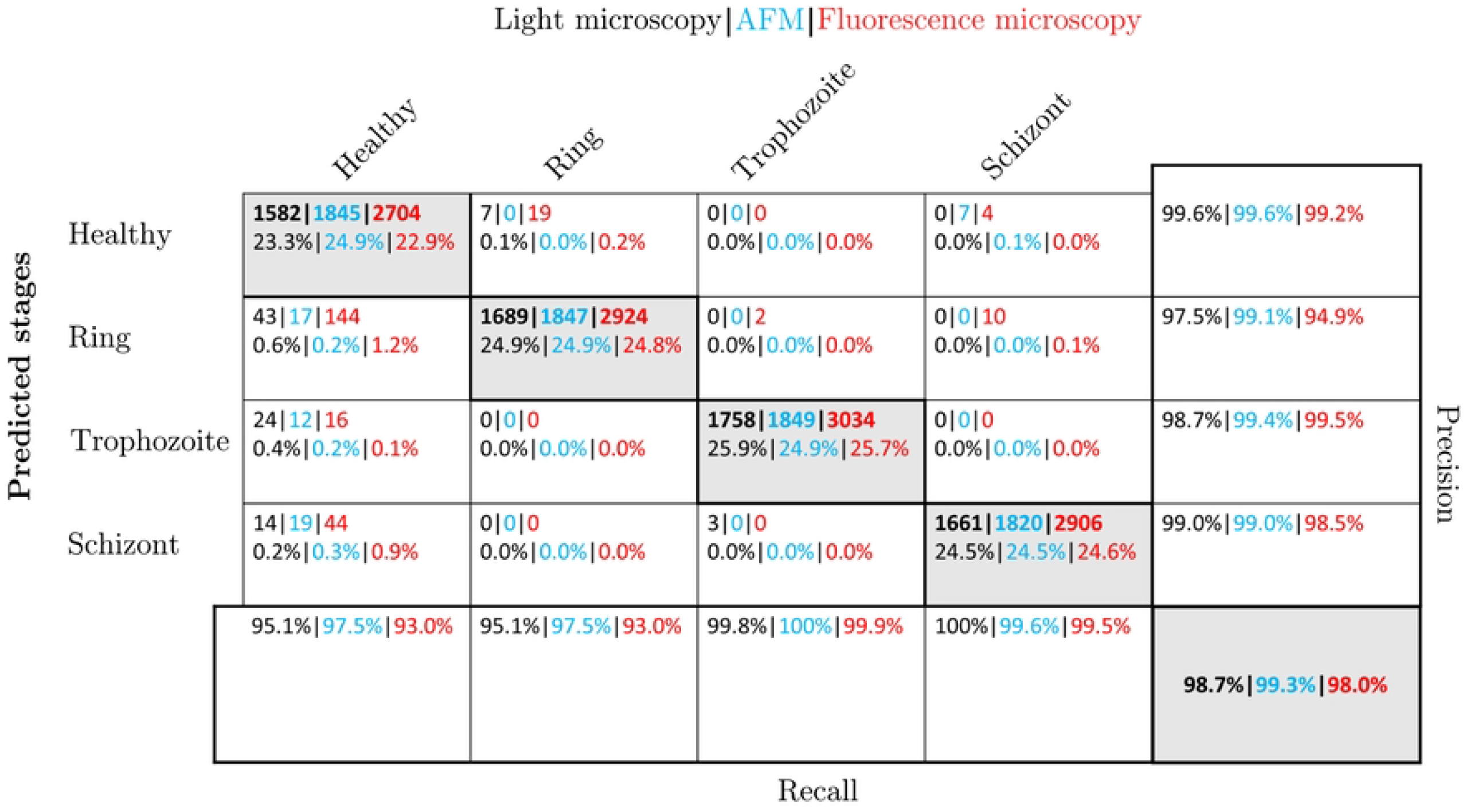
Show image. The input file is shown as image.

If the image was recorded by AFM, the next step requires thresholding to enhance its contrast. While the value can be adapted by the user, the algorithm suggests a pre-calculated number, as shown Fig 9. In case of the light microscopy techniques, this step is skipped.

**Figure 9.**
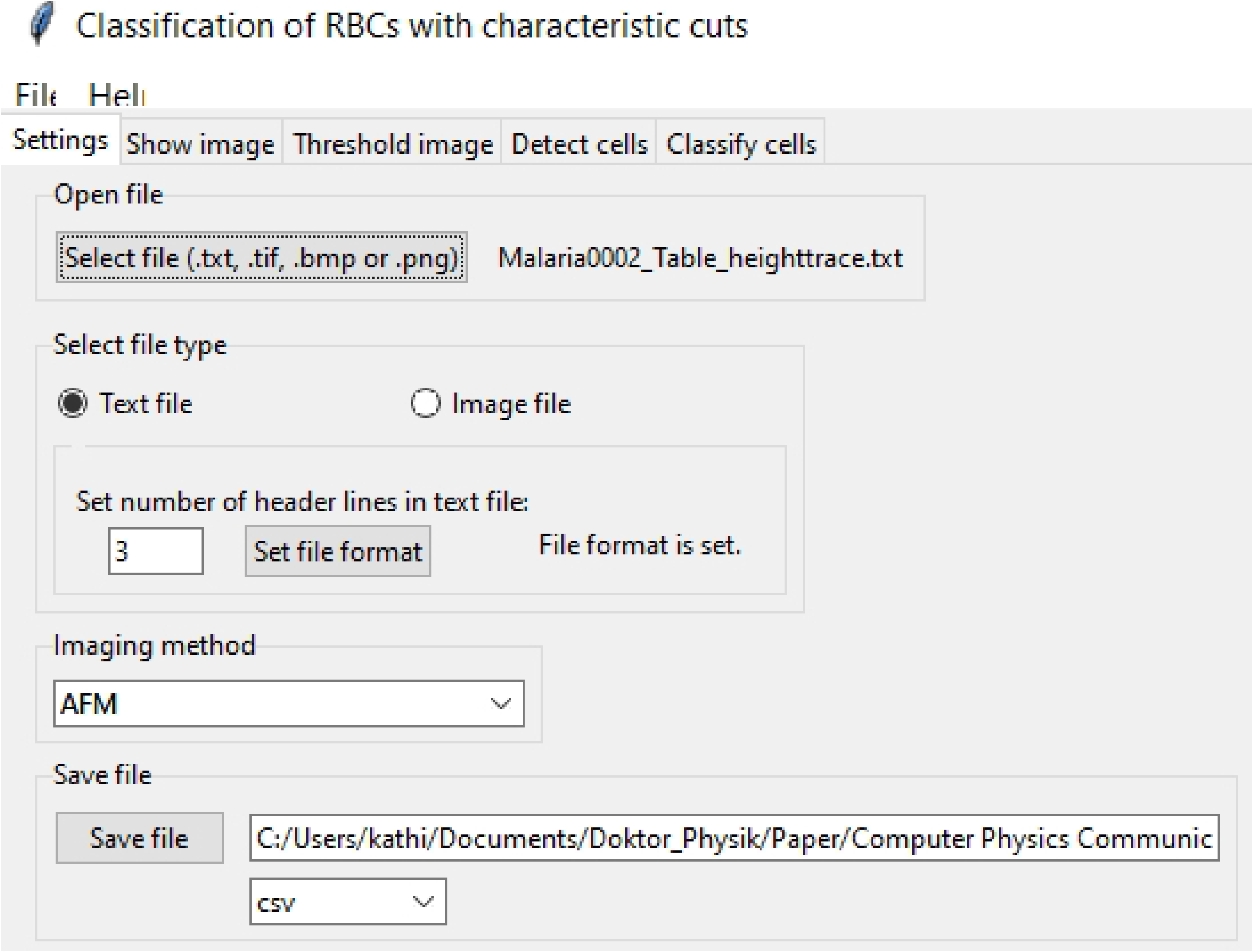
Threshold image. This tab provides the option to enhance the image contrast by thresholding (disabled for light and fluorescence microscopy)

The next tab triggers the cell detection algorithm. Depending on the size, contrast, and brightness of the image, the algorithm offers the option to control the detection by manually changing the calculation parameters and further allows contrast enhancement. Optionally, the parameters can be set back to the default value. During this step, the dimension of the input data is reduced to the two characteristic cuts to capture the strongest features associated with the presence or absence and the stage of the malaria parasites, see Fig 10.

**Figure 10.**
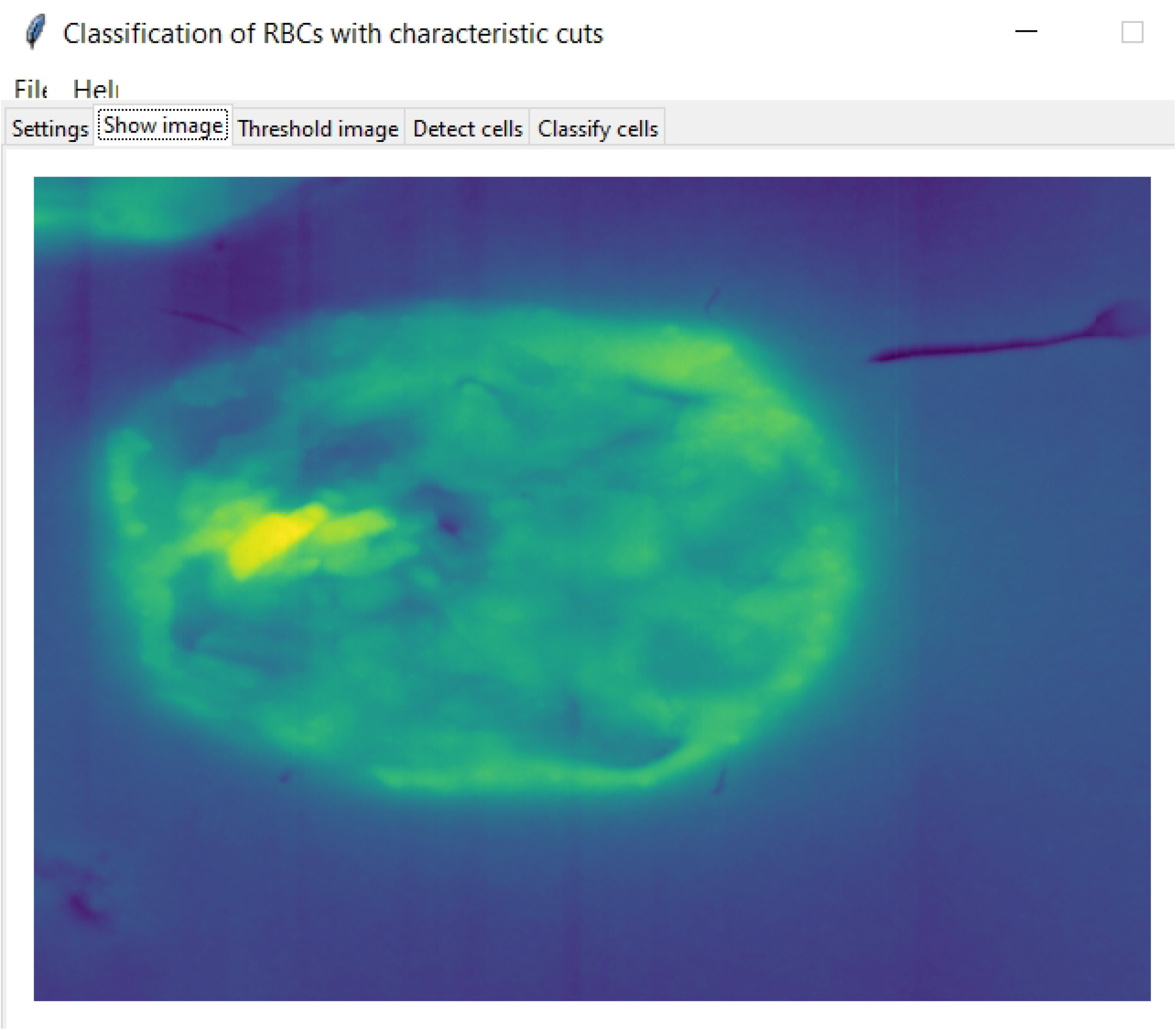
Detect cells. RBCs in the images are detected by the cell detection algorithm. To fine-tune, the algorithm offers the possibility to enhance the image and to set the parameters for the detection manually.

In the final part of the program, the user can load the pre-trained NNs and start the stage-specific prediction of the detected RBCs. This triggers the option to change false predictions accordingly. The algorithm further provides the possibility to add new data and to retrain each NN. The last step then returns the statistics of the analysed RBCs in form of a text file or table. The corresponding tab is shown in Fig 11.

**Figure 11.** Cell classification. The intra-erythrocytic stages are predicted and can be altered manually. Optionally, the neural network can be retrained with new data.

## Data Availability

All data produced are available online at

https://zenodo.org/record/6866337

https://github.com/KatharinaPreissinger/Malaria_stage_classifier

## Supplementary data

S1 Data. This ZIP file contains the code, the pretrained neural networks, the sample images, a README file, the logo for the interface, and sample images for testing purposes. The file is available at https://github.com/KatharinaPreissinger/malaria_stage_classifier.

S2 Data. Documentation: This is the documentation of the Malaria Stage Classifier. The current version is available at https://malaria-stage-classifier.readthedocs.io/en/latest/index.html

S3 Data. Training, validation, and test set: This is the data, which can be used to retrain the neural network with new cell images. The dataset is available at https://zenodo.org/record/6866337.

## Acknowledgments

This research was supported by the National Research, Development and Innovation Office of Hungary (K119493, VEKOP-2.3.2-16-2017-00013, NKP-2018-1.2.1-NKP-2018-00005), the BME-Biotechnology FIKP grant of EMMI (BME FIKP-BIO), the BME-Nanotechnology and Materials Science FIKP grant of EMMI (BME FIKP-NAT), the National Heart Programme (NVKP-16-1-2016-0017), K124966 and K135360, National Bionics Programme ED 17-1-2017-0009 and SE FIKP-Therapy Grant.

## Author contributions

Conceptualisation: Katharina Preißinger, János Török Investigation: Katharina Preißinger Software: Katharina Preißinger Supervision: István Kézsmárki Validation: Katharina Preißinger Visualisation: Katharina Preißinger Writing - original draft: Katharina Preißinger

## Notes

### Competing Interest Statement

The authors have declared no competing interest.

## References

1. T. A. Ghebreyesus, World malaria report 2021, Tech. rep., World health organisation (2021).

2. P. Hough, Machine analysis of bubble chamber pictures, in: Proc. Int. Conf. High Energy Accelerators and Instrumentation, 1959, p. 5.

3. K. Preißinger, I. Kézsmárki, J. Török, Reducing data dimension boosts neural network-based stage-specific malaria detection, Scientific Reports 12 (1) (2022) 1–14 doi:10.1038/s41598-022-19601-x.

4. S. Harvey, S. Incardona, N. Martin, Quality issues with malaria rapid diagnostic test accessories and buffer packaging: findings from 5-country private sector project in Africa, Malaria Journal 16 (160) (2017).

5. K. Preißinger, P. Molnar, B. Vertessy, I. Kezsmarki, M. Kellermayer, Stage-Dependent Topographical and Optical Properties of Plasmodium Falciparum-Infected Red Blood Cells, Journal of Biotechnology and Biomedicine 4 (3) (2021) 132–146. doi:10.26502/jbb.2642-91280040.

6. P. Lebel, R. Dial, V. N. P. Vemuri, V. Garcia, Label-free imaging and classification of live P. falciparum enables high performance parasitemia quantification without fixation or staining, Plos computational biology 17 (8) (2021) 1–29. doi:10.1371/journal.pcbi.1009257. URL https://doi.org/10.1371/journal.pcbi.1009257%0AEditor:

7. A. Mehrjou, Automatic Malaria Diagnosis System, International Conference on Robotics and Mechatronics 2013 (2013) 205–211.

8. M. Poostchi, K. Silamut, R. J. Maude, S. Jaeger, G. Thoma, O. Tropical, G. Health, Image analysis and machine learning for detecting malaria, Translational Researcch 194 (2019) 36–55. doi:10.1016/j.trsl.2017.12.004.Image.

9. M. C. Mushabe, R. Dendere, T. S. Douglas, Automated detection of malaria in Giemsa-stained thin blood smears, Annual international conference of the IEEE engineering in medicine and biology society 2013 (3698-701) (2013).

10. K. Torres, C. M. Bachman, C. B. Delahunt, J. Alarcon Baldeon, F. Alava, D. Gamboa Vilela, S. Proux, C. Mehanian, S. K. McGuire, C. M. Thompson, T. Ostbye, L. Hu, M. S. Jaiswal, V. M. Hunt, D. Bell, Automated microscopy for routine malaria diagnosis: A field comparison on Giemsa-stained blood films in Peru, Malaria Journal 17 (1) (2018) 1–11. doi:10.1186/s12936-018-2493-0. URL https://doi.org/10.1186/s12936-018-2493-0

11. I. R. Dave, K. P. Upla, Computer aided diagnosis of Malaria disease for thin and thick blood smear microscopic images, 2017 4th International Conference on Signal Processing and Integrated Networks, SPIN 2017 2017 (2017) 561–565. doi:10.1109/SPIN.2017.8050013.

12. E. Nagao, O. Kaneko, J. A. Dvorak, Plasmodium falciparum-Infected Erythrocytes : Qualitative and Quantitative Analyses of Parasite-Induced Knobs by Atomic Force Microscopy, Journal of Structural Biology 44 (2000) 34–44. doi:10.1006/jsbi.2000.4236.

13. H. Shi, Z. Liu, A. Li, J. Yin, A. G. Chong, K. S. Tan, Y. Zhang, C. T. Lim, Life Cycle-Dependent Cytoskeletal Modifications in Plasmodium falciparum Infected Erythrocytes, PLoS ONE 8 (4) (2013) 1–10. doi:10.1371/journal.pone.0061170.

14. D. Perez-guaita, K. Kochan, M. Batty, C. Doerig, J. Garcia-bustos, S. Espinoza, D. Mcnaughton, P. Heraud, B. R. Wood, Multispectral Atomic Force Microscopy-Infrared Nano-Imaging of Malaria Infected Red Blood Cells, Analytical Chemistry 90 (2018) 3140–3148. doi:10.1021/acs.analchem.7b04318.

15. K. E. delas Penas, P. T. Rivera, P. C. Naval, Malaria Parasite Detection and Species Identification on Thin Blood Smears using a Convolutional Neural Network, in: International Conference on Connected Health, 2017. doi:10.1109/CHASE.2017.51.

16. Q. Ammar, A. Mohsen, A. S.-u. Hassan, C. Chen, A. Imran, G. Rasul, A dataset and benchmark for malaria life-cycle classification in thin blood smear images, Neural Computing and Applications 6 (2021). doi:10.1007/s00521-021-06602-6. URL https://doi.org/10.1007/s00521-021-06602-6

17. V. V. Makkapati, R. M. Rao, Segmentation of malaria parasites in peripheral blood smear images, in: 2009 IEEE International Conference on Acoustics, Speech and Signal Processing, IEEE, 2009, pp. 1361–1364.

18. D. Das, M. Ghosh, Probabilistic Prediction of Malaria using Morphological and Textural Information, in: 2011 International Conference on Image Information Processing (ICIIP 2011), IEEE, 2011.

19. A. Pinkaew, T. Limpiti, A. Trirat, I. Vxe, L. Ylyd, V. X. E. Lpdjhv, Automated classification of malaria parasite species on thick blood film using support vector machine, in: Th 2015 Biomedical Engineering International Conference, IEEE, 2015, pp. 8–12.

20. M. S. Davidson, C. Andradi-brown, S. Yahiya, J. Chmielewski, A. J. O. Donnell, P. Gurung, M. D. Jeninga, P. Prommana, D. W. Andrew, M. Petter, C. Uthaipibull, M. J. Boyle, G. W. Ashdown, J. D. Dvorin, S. E. Reece, D. W. Wilson, K. A. Cunningham, D. M. Ando, M. Dimon, J. Baum, Automated detection and staging of malaria parasites from cytological smears using convolutional neural networks, Biological Imaging 1 (e2) (2021) 1–13. doi:10.1017/S2633903X21000015.

21. A. Rahman, H. Zunair, M. S. Rahman, J. Q. Yuki, Improving Malaria Parasite Detection from Red Blood Cell using Deep Convolutional Neural Networks, arXiv 1907.10418 (2019) 1–33.

22. K. Sriporn, C.-F. Tsai, C.-E. Tsai, P. Wang, Analyzing Malaria Disease Using Effective Deep Learning Approach, Diagnostics 10 (10) (2020) 1–22.

23. D. O. Oyewola, E. G. Dada, S. Misra, R. Damaševičius, A Novel Data Augmentation Convolutional Neural Network for Detecting Malaria Parasite in Blood Smear Images, Applied Artificial Intelligence 2022 (2022) 1–22. doi:10.1080/08839514.2022.2033473.

24. J. Brownlee, Introduction to Dimensionality Reduction for Machine Learning (2020). URL https://machinelearningmastery.com/dimensionality-reduction-for-machine-learning/

25. S. Kareem, I. Kale, R. C. Morling, Automated malaria parasite detection in thin blood films:-A hybrid illumination and color constancy insensitive, morphological approach, IEEE Asia-Pacific Conference on Circuits and Systems, Proceedings, APCCAS 2012 (2012) 240–243. doi:10.1109/APCCAS.2012.6419016.

26. C. Shorten, T. M. Khoshgoftaar, A survey on Image Data Augmentation for Deep Learning, Journal of Big Data 6 (1) (2019). doi:10.1186/s40537-019-0197-0. URL https://doi.org/10.1186/s40537-019-0197-0

27. N. Otsu, A threshold selection method from gray-level histograms, IEEE Transactions on Systems, Man, and Cybernetics SMC-9 (1) (1979) 62–66.

28. F. Lundh, A. Clark, Pillow (PIL Fork) Reference (2022). URL https://pillow.readthedocs.io/en/stable/reference/index.html

29. Tutorials OpenCV, Hough Circle Transform (2021). URL https://docs.opencv.org/3.4/d4/d70/tutorial_hough_circle.html

30. J. Canny, A Computational Approach to Edge Detection, IEEE Transactions on Pattern Analysis and Machine Intelligence PAMI-8 (6) (1986) 679–698. doi:10.1109/TPAMI.1986.4767851.

31. D. H. Hubel, T. N. Wiesel, Receptive fields and functional architecture of monkey striate cortex, The Journal of Physiology 195 (1) (1968) 215–243. doi:10.1113/jphysiol.1968.sp008455.

32. S. Abbas, T. Djkstra, Detection and stage classification of Plasmodium falciparum from images of Giemsa stained thin blood films using random forest classifiers, Diagnostic Pathology 15 (130) (2020) 1–11.

33. H. Prasanna, S. R. Mahadeva; Yegnanarayana, B.; Pinto, Joel Praveen; Hermansky, Analysis of Confusion Matrix to Combine Evidence for Phoneme Recognition, Tech. rep., IDIAP Research institute Switzerland (2007).

34. T. Oliphant, J. k. Millma, A guide to NumPy, Trelgol Publishing (2006). doi:DOI:10.1109/MCSE.2007.58.

35. W. McKinney, Data Structures for Statistical Computing in Python, Proceedings of the 9th Python in Science Conference 1 (Scipy) (2010) 56–61. doi:10.25080/majora-92bf1922-00a.

36. OpenCV Team, OpenCV (2022). URL https://opencv.org/about/

37. S. image development team, Scikit-image: image processing in python (2022). URL https://scikit-image.org/docs/stable/user_guide.html

38. M. development team, Matplotlib 3.5.2 documentation (2022). URL https://matplotlib.org/stable/index.html

39. NVIDIA, TensorFlow (2022). URL https://docs.nvidia.com/deeplearning/frameworks/tensorflow-user-guide/index.html

40. Python Software Foundation, Graphical user interfaces with Tk (2022). URL https://docs.python.org/3/library/tk.html

